# Electroconvulsive Therapy Modulates Loudness Dependence of Auditory Evoked Potentials: A Pilot MEG Study

**DOI:** 10.1101/2024.04.26.24306462

**Authors:** Michael Dib, Jeffrey D. Lewine, Christopher C. Abbott, Zhi-De Deng

**Author notes:** Correspondence: Zhi-De Deng. These authors share senior authorship (Christopher C. Abbott, Zhi-De Deng).

## Abstract

Electroconvulsive therapy (ECT) remains a critical intervention for treatment-resistant depression (MDD), yet its neurobiological underpinnings are not fully understood. This pilot study utilizes high-resolution magnetoencephalography (MEG) in nine depressed patients receiving right unilateral ECT, to investigate the changes in loudness dependence of auditory evoked potentials (LDAEP), a proposed biomarker of serotonergic activity, following ECT. We hypothesized that ECT would reduce the LDAEP slope, reflecting enhanced serotonergic neurotransmission. Contrary to this, our findings indicated a significant increase in LDAEP post-ECT (*t*_8_ = 3.17, *p* = .013). The increase in LDAEP was not associated with changes in depression severity or cognitive performance, as assessed by the Hamilton Depression Rating Scale (HAMD-24) and Repeatable Battery for the Assessment of Neuropsychological Status (RBANS). We discussed potential mechanisms for the observed increase, including ECT’s impact on serotonergic, dopaminergic, glutamatergic, and GABAergic receptor activity, neuroplasticity involving brain-derived neurotrophic factor (BDNF), and inflammation modulators such as TNF-*alpha*. Our results suggest a complex interaction between ECT and these neurobiological systems, rather than a direct reflection of serotonergic neurotransmission.

## 1 INTRODUCTION

Major depressive disorder (MDD) remains a pervasive global health challenge, affecting millions worldwide and ranking among the leading causes of disability. MDD leads to substantial healthcare costs and contributing heavily to the overall disease burden (1). Despite the widespread use of antidepressant medications, many patients do not achieve sustained relief. As an alternative, neuromodulation therapies such as electroconvulsive therapy (ECT) play a vital role. ECT is a well-established intervention that demonstrates exceptional efficacy in multiple psychiatric disorders, including MDD. It involves the administration of electrical currents, either unilateral or bilateral electrode placements on the patient’s head, to induce a brief, controlled seizure. Ultimately, this process is thought to elicit reorganization of key cortical networks involved with mood and cognition. However, ECT can cause significant adverse effects, such as memory loss and confusion, rendering ECT to be reserved for severely treatment resistant patients (2, 3). entifying the specific neurophysiological changes induced by ECT could lead to the development of safer and more effective treatments. While the optimal stimulation methods and parameters are still being investigated, ECT remains essential for managing treatment-resistant depression.

More than eight decades have passed since its introduction as a clinical intervention, the precise neurobiological mechanisms underpinning ECT’s therapeutic effect remain elusive. Current research suggests that ECT’s benefits are likely achieved through multiple mechanisms. These include, but are not limited to, changes in neurotransmitter transmission, enhancement of neurotrophic and neuroplastic activities, modulation of cortical networks, reduction of neuroinflammation, and regulation of the endocrine system. (3, 4, 5, 6, 7, 8, 9, 10). Given the historical precedence of the monoaminergic theory of depression, a plethora of studies in both animals and humans have sought out to determine whether ECT’s efficacy is related to changes in serotonergic activity (11, 12). While the evidence remains inconclusive, ECT appears to have some notable effect on serotonergic neurotransmission (3). A significant challenge in this area is that peripheral biomarker measurements do not reliably reflect neurotransmitter levels in the brain. Advanced neuroimaging techniques can be employed to gather insight into the effect of treatments such as ECT on neurotransmitter activity.

Loudness dependence of auditory evoked potentials (LDAEP) is a method used to measure the response of cortical potentials to variations in the intensity (i.e., loudness) of auditory stimuli. The relationship between stimuli loudness and evoked potential amplitudes in the primary auditory cortex has been suggested as an indicator of serotonergic neurotransmission (13). Initial studies in animal models reported that higher serotonin activity was correlated with less dependence on stimulus intensity (i.e., similar amplitudes in cortical evoked potentials regardless of loudness). Conversely, lower serotonin activity was correlated with loudness-dependent changes in evoked potential amplitudes (13, 14). Furthermore, LDAEP has been proposed to be a protective mechanism in auditory processing, which helps prevent overstimulation and excitotoxicity (15). Within the primary auditory cortex, the neurobiological mechanisms of a reduced LDAEP being associated with high serotonin activity is proposed to rely on serotonergic modulation of cortical excitability. This modulation occurs indirectly via GABA-ergic interneurons, which express excitatory 5-HT2A receptors, and directly via pyramidal cells, which express both excitatory 5-HT2A and inhibitory 5-HT1A receptors (13, 14, 16, 17).

An abundance of literature now exists exploring the relationship between the LDAEP and other neurotransmitter systems and biomarkers (18, 19). While evidence for the LDAEP’s relationship with serotonin is controversial, it is clear serotonergic activity serves a critical role in the functioning of the primary auditory cortex (18, 20, 21, 22, 23, 24). Numerous studies provide robust support for the influence of serotonergic activity on neuronal functioning across auditory processing pathways (25, 26, 27, 28). Notably, a recent positron emission tomography (PET) study on the molecular mechanisms underlying the LDAEP reported that this biomarker is strongly and positively correlated with 5-HT1A binding in the temporal cortex, specifically in the location of the primary auditory cortex (29).

Additionally, studies have demonstrated that serotonin plays a crucial role at the intersection of psychiatric disorders and auditory conditions, including tinnitus and hearing loss (25, 26). In particular, MDD has been reported to be associated with impaired auditory processing. Studies indicate that deviations in serotonergic activity are evident in the auditory cortices of individuals with depression compared to controls (23, 30, 31, 32, 33). For example, increased 5-HT1A binding and decreased 5-HT2A binding specifically within the primary auditory cortex in depressed patients has been reported (23). Moreover, treatments for depression, including ECT, have been shown to have a significant effect on auditory processing, demonstrated via increased activity, excitability, and intrinsic connectivity within the auditory cortices (30, 31, 34, 35, 36). Additionally, ECT has shown a pronounced impact on auditory evoked potentials, further underscoring the complex interplay between serotonergic modulation and auditory functions in psychiatric contexts (37, 38).

Given the efficacy of serotonergic agents such as selective serotonin reuptake inhibitors, and more recently psychedelics like psilocybin, in the treatment of depression, it is likely regulation of this monoamine system serves a pivotal role in ECT’s efficacy (39). Studies show mixed results regarding ECT’s impact on serotonergic receptors. For instance, some reports indicate that electroconvulsive stimuli result in decreased binding and activity of both 5-HT1A and 5-HT2A receptors (12, 40, 41, 42). However, other studies reveal no change in 5-HT1A activity, an increase in 5-HT2 activity, and enhanced serotonin transporter (SERT) receptor levels following ECT (43, 44, 45, 46). Despite these discrepancies, there is broad consensus that ECT has a robust impact on serotonergic receptors in the treatment of multiple psychiatric disorders (8, 12, 47, 48).

This study utilizes high-resolution magnetoencephalography (MEG) to measure cortical activity and determine the LDAEP in individuals before and after ECT. The primary aim is to explore the changes in the central serotonergic neurotransmission attributable to ECT, by analyzing variations in LDAEP. We hypothesize that ECT will decrease LDAEP, indicative of enhanced serotonergic neurotransmission. This approach not only promises to deepen our understanding of the neurochemical environment in patients undergoing ECT but also sheds light on the neurobiological mechanisms that underpin ECT’s effectiveness.

## 2 METHODS

Study participants and ECT treatment Ethical approval was obtained from the Human Research Protections Office at the University of New Mexico (UNM) before study initiation. The research was conducted in full compliance with the ethical standards outlined in the Declaration of Helsinki. Patients were recruited from the UNM Mental Health Center’s inpatient and outpatient services. All patients either had the decisional capacity to consent or, where necessary, provided assent with a surrogate decision-maker giving formal consent. Prior to receiving treatment, the average score on the 24-item Hamilton Depression Rating Scale (HAMD-24) for these patients was 37.2 (12.8). Additionally, the mean score on the total Repeatable Battery for the Assessment of Neuropsychological Status (RBANS) was 82.9 (19.4). All patients completed a full course of electroconvulsive therapy (ECT) using the ultra-brief pulse width, right unilateral electrode placement as previously described (49). During the initial session, the seizure threshold was determined using a dose titration method, which then guided the dosage for subsequent treatments. Specifically, the stimulus dosage was set at six times the threshold. Treatments were administered thrice weekly, and continued until an adequate clinical response was achieved or a decision was made to cease treatment due to non-response.

### 2.1 MRI

All MRI scans were conducted using the 3-Tesla Siemens Trio scanner at the Mind Research Network (MRN). High-resolution T1-weighted structural images were acquired using a 5-echo MPRAGE sequence with the following parameters: echo times (TE) of 1.64, 3.5, 5.36, 7.22, and 9.08 ms; repetition time (TR) of 2.53 s; inversion time (TI) of 1.2 s; a flip angle of 7°; a single excitation; a slice thickness of 1 mm; a field of view of 256 mm; and a resolution of 256 × 256. Structural MRI preprocessing and the delineation of structural images were conducted using FreeSurfer 4.5.0 software (https://surfer.nmr.mgh.harvard.edu) (50).

### 2.2 MEG acquisition and data processing

Prior to and following the ECT course, patients underwent MEG scans (see Figure 1). MEG recordings were captured using the Elektra Neuromag VectorView 306 system, which is equipped with 102 magnetometers and 204 planar gradiometers. To ensure accurate alignment, the MRIs were coregistered with scalp fiducial markers. While seated inside the MEG helmet, patients were exposed to a series of auditory tones at five different intensity levels: 55, 65, 75, 85, and 95 dB. The tones were emitted through biauricular earbuds at a frequency of 2 kHz, lasting 50 ms each. The tones were presented in a random sequence, with each intensity block comprising 22 tones, resulting in a total of 110 trials per intensity level.

**Figure 1.**
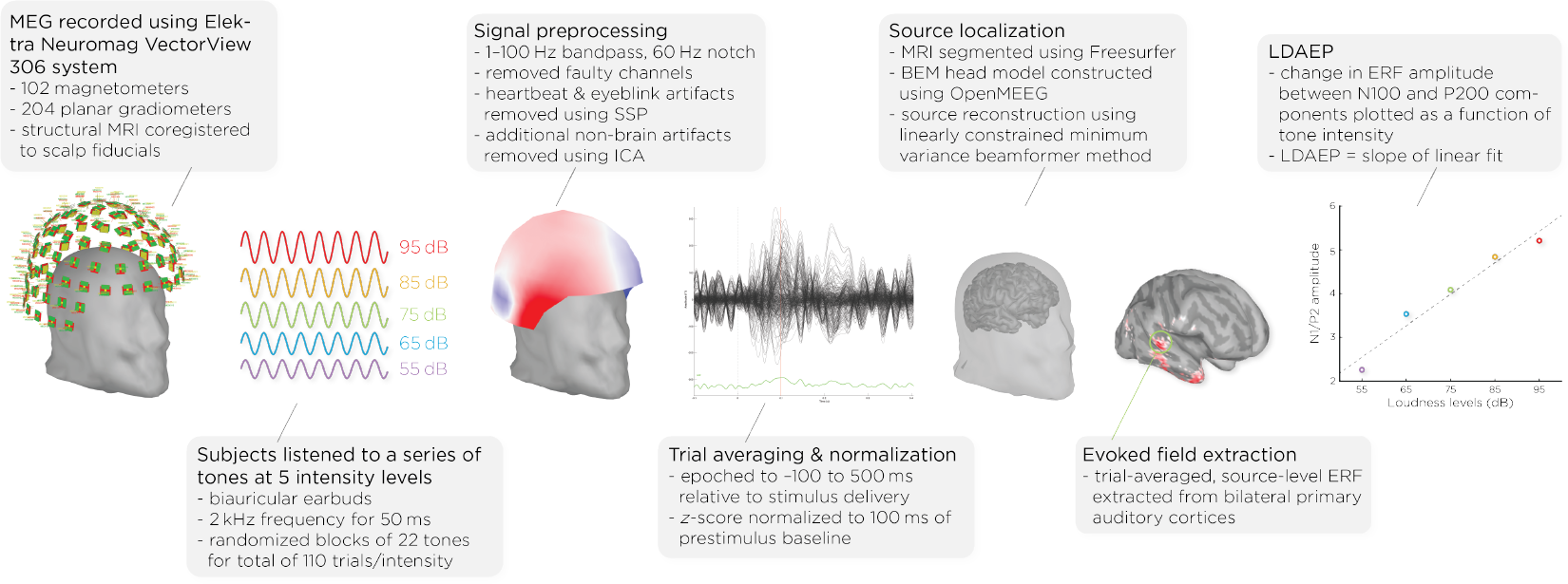
Workflow of MEG acquistion and data processing, detailing the steps from signal acquisition to data analysis. It shows the MEG setup, signal preprocessing, auditory stimulus presentation, data normalization, brain activity source localization, and the quantification of auditory evoked potentials (AEPs) across different sound intensity levels.

Data analysis was performed with Brainstorm 3 (51), which is documented and freely available for download online under the GNU general public license (http://neuroimage.usc.edu/brainstorm). The MEG data was filtered using a 1–100 Hz bandpass filter and a 60 Hz notch filter to eliminate electrical line noise. Malfunctioning channels were identified and excluded. Artifacts arising from cardiac activity and eye blinks were removed via signal-space projection, and independent component analysis was used to eliminate other non-brain artifacts. Auditory events were defined for a time window from −100 to 500 ms around the tone’s presentation. The data was normalized using the Z-transformation relative to the 100 ms pre-stimulus baseline.

Cortical structures were derived from each subject’s MRI scans using FreeSurfer, and aligned with a standard brain atlas for cortical reconstruction. The head model for the forward model utilized the symmetric boundary element method (BEM) implemented in OpenMEEG, provided by the INRIA institute. This model established a computational link between the neuronal activity in the source space and the recorded MEG data in the sensor space, considering the conductive properties of head tissues. The inverse model, which infers neural activity from the MEG data, was computed using a data covariance matrix through the linearly constrained minimum variance (LCMV) beamforming technique, focusing on auditory evoked potentials. Trial-averaged, source-level event-related fields (ERFs) were extracted from the bilateral primary auditory cortices. Finally, the LDAEP was calculated by evaluating the change in normalized ERF amplitude between the N100 and P200 components from the trial-averaged epochs. The LDAEP is represented by the slope of the linear regression line fitted to these data points.

### 2.3 Statistical analysis

We evaluated the distribution of our data for normality using the Shapiro-Wilk test. The tests indicated normality in the changes in the LDAEP slope (*p* = 0.098), HAMD scores (*p* = 0.263), and total RBANS scores (*p* = 0.105). To investigate longitudinal differences in LDAEP slopes, depression scores, and cognitive functioning scores, we employed paired *t*-tests. Additionally, we explored correlations among the pre-treatment and post-treatment LDAEP slopes, the degree of their changes, and the baseline, post-treatment, and changes in HAMD-24 and RBANS scores.

## 3 RESULTS

### 3.1 Demographics and clinical outcomes

Six of nine patients responded (> 50% reduction in HAMD-24 from baseline) to ECT with an average post-ECT HAMD-24 score of 9.1 ± 7.6 (*t*_8_ = 5.60, *p* < .001). We collected seven of the nine patients’ RBANS data, and found, on average, their cognitive functioning did not change with ECT (*t*_6_ = 0.358, *p* = .73). Demographics and clinical measures before and after ECT treatment are summarized in Table 1.

**Table 1.** Demographics and clinical measures before and after ECT treatment.

| Measure | Pre-ECT (Mean ± SD) | Post-ECT (Mean ± SD) | $t$ | $p$-value |
| --- | --- | --- | --- | --- |
| $N$ (no. female) | 9 (6) | - | - | |
| Age, years | $68.1 \pm 10.7$ | - | - | |
| LDAEP | $0.41 \pm 0.64$ | $0.75 \pm 0.74$ | 3.17 | .013 |
| HAMD-24 | $37.2 \pm 12.8$ | $9.1 \pm 7.6$ | 5.60 | $< 0.001$ |
| RBANS Scores |  |  |  |  |
| Total | $82.9 \pm 19.4$ | $84.4 \pm 18.0$ | 0.36 | .73 |
| Immediate memory | $78.0 \pm 25.4$ | $83.1 \pm 24.2$ | 0.84 | .44 |
| Visuospatial/Constructional | $84.7 \pm 26.1$ | $90.7 \pm 17.7$ | 0.79 | .46 |
| Language | $92.6 \pm 8.9$ | $89.7 \pm 8.0$ | 0.79 | .46 |
| Attention | $92.4 \pm 14.7$ | $89.6 \pm 17.9$ | 1.05 | .34 |
| Delayed memory | $85.3 \pm 22.1$ | $85.7 \pm 22.2$ | 0.08 | .94 |

### 3.2 Change in LDAEP

Figure 2 shows the auditory evoked potentials before and after ECT treatment, with responses at varying stimulus loudness levels. The LDAEP slope significantly increased following ECT treatment from 0.41 ± 0.64 to 0.75 ± 0.74) (Cohen’s *d* = 0.49, *t*_8_ = 3.17, *p* = .013) (Figure 3).

**Figure 2.**
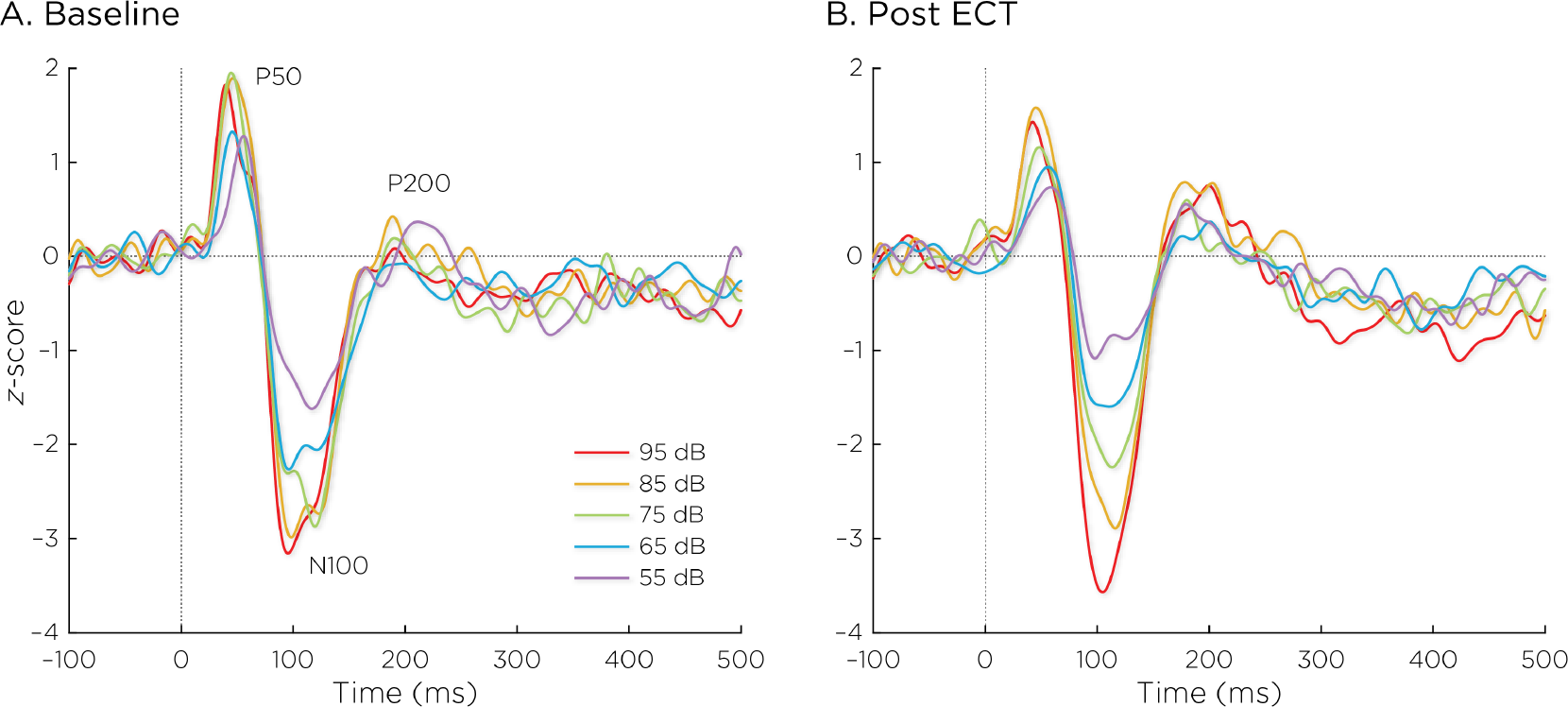
Auditory evoked potentials (A) before and (B) after ECT treatment, with responses at sound pressure levels of 55–95 dB. The change in normalized AEP amplitude between the N100 and P200 (N1/P2) components of the trial-averaged epochs was calculated. The LDAEP is calculated as the slope of linear regression line that best fits the N1/P2 amplitudes at each sound pressure level.

**Figure 3.**
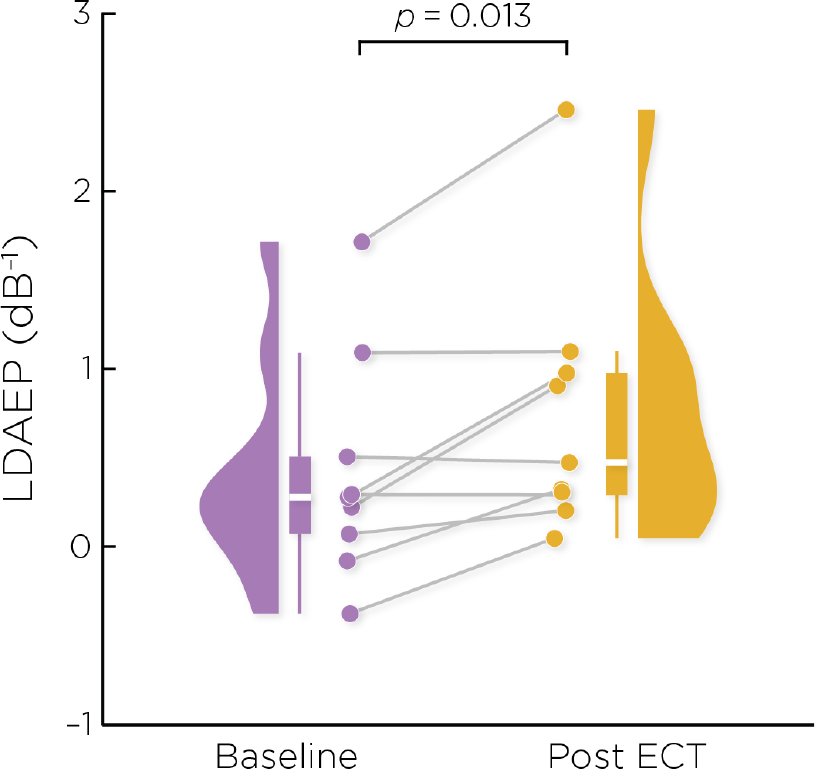
Individual changes in the pre- and post-ECT LDAEP slope measure. There was a significant increase post-ECT compared to baseline (Cohen’s *d* = 0.49, *t*_8_ = 3.17, *p* = .013).

### 3.3 Correlations between LDAEP and depression and cognition scores

The change in the LDAEP slope was not significantly correlated with baseline (*r* = −0.079, *p* = .84), post-treatment (*r* = 0.066, *p* = .87), or changes in HAMD-24 scores (*r* = 0.101, *p* = .80). The pre-ECT LDAEP slope was not significantly correlated with baseline (*r* = −0.194, *p* = .62) nor changes in HAMD-24 scores (*r* = 0.339, *p* = .37).

We focused on assessing correlations with RBANS total scores. The change in LDAEP slope was not significantly correlated with the baseline (*r* = 0.208, *p* = .66), post-treatment (*r* = 0.403, *p* = .37), or changes (*r* = 0.276, *p* = .55) in the RBANS total score. The pre-ECT LDAEP slope was significantly correlated with the baseline RBANS total score (*r* = 0.855, *p* = .014), but not correlated with changes in the RBANS total score (*r* = −0.352, *p* = .44).

## 4 DISCUSSION

In this study, we used LDAEP as a cortical activity biomarker to monitor changes in neurotransmitter activity induced by ECT. Our initial hypothesis posited that ECT would mitigate symptoms of depression by boosting serotonergic neurotransmission, which would manifest as a reduced LDAEP, reflected by weakening of the response amplitude as a function of sound intensity levels. However, our findings revealed a significant increase in LDAEP post-ECT. Interestingly, the alterations in LDAEP did not correlate with changes in depression severity or cognitive performance.

The neurochemical underpinnings of LDAEP suggest that ECT should lead to a reduction in serotonergic tone within the primary auditory cortex, but this assumption is subject to debate. Studies on the relationship between LDAEP and serotonergic activity have harbored conflicting evidence and perspectives (19, 24, 52, 53). While numerous studies have found that the LDAEP is sensitive to acute changes in serotonergic activity, such as following administration of serotonergic-reuptake inhibitors, other studies have presented contrasting findings (54, 55, 56, 57, 58, 59, 60). A recent narrative review by Kangas et. al stated that LDAEP studies have generally yielded no consistent difference between depressed and non-depressed controls, though there appears to be a relationship with depression-subtypes (18). Given this context, a possible explanation for our results not supporting the original hypothesis could be that the LDAEP does not precisely mirror serotonergic tone in the primary auditory cortex.

Our findings did reveal a significant modulation of the LDAEP after ECT treatment. Intriguingly, the observed increase in LDAEP may indicate that ECT prompted a reduction in serotonergic receptor activity within the primary auditory cortex, particularly 5-HT1A and 5-HT2 receptors. This algins with multiple studies demonstrating a reduction in 5-HT2 and 5-HT1A receptors in humans and non-human primates following ECT (40, 41, 42, 61, 62). The initial LDAEP studies in animal models bolster the plausibility of ECT-induced reduction in serotonergic receptors, in that a 5-HT1A agonist decreased the LDAEP and a 5-HT2A antagonist increased the LDAEP, suggesting that decreased serotonin receptor activity results in a strengthened LDAEP (13, 63). However, pre-clinical studies have largely found an upregulation of 5-HT1A and 5-HT2A receptors after electroconvulsive stimuli in animal models (44, 46, 64). This discrepancy underscores the need for further research to understand ECT’s impact on serotonergic receptor activity in humans more comprehensively.

Several alternative explanations as to why the LDAEP increased following ECT may be plausible. The LDAEP has been shown to covary with symptom severity in disorders such as ADHD, schizophrenia, and Parkinson’s disease, all of which are strongly linked to dopaminergic dysregulation (65, 66, 67). Given that ECT has been found to significantly alter dopaminergic neurotransmission (44, 68, 69, 70, 71), one possible explanation for the heightened LDAEP could be ECT’s direct influence on dopamine receptor and transporter activities. Furthermore, considering that the LDAEP results from both excitatory and inhibitory post-synaptic potentials within the primary auditory cortex, the altered LDAEP could reflect changes in glutamatergic and GABAergic functions. Indeed, ECT has been shown to increase GABA concentration, normalize glutamate deficits, and alter excitation/inhibition ratios (26, 72, 73, 74, 75). For instance, the administration of a glutamatergic NMDA antagonist has been reported to blunt LDAEP, suggesting that increased glutamatergic activity correlates with a heightened LDAEP (76). On the other hand, a study reported that the LDAEP is not associated with GABA levels (77). Further exploration of the relationships between ECT, auditory cortical activity, and excitatory and inhibitory neurotransmitters could yield valualbe insights.

Beyond neurotransmitter effects, there is a consistent body of evidence indicating that ECT is associated with increased gray matter volume in the temporal lobes, including the superior temporal gyrus (5, 78, 79, 80), where the primary auditory cortex is situated. It is possible that neurotrophic effects are related to the increased LDAEP following ECT in this study. A recent meta-analysis concluded that ECT directly increases concentrations of brain-derived neurotrophic factor (BDNF) (81). Similarly, the LDAEP has been found to be significantly positively correlated with serum BDNF levels (82). One possibility is that ECT’s robust neuroplastic effects within the temporal lobe are related to the modulation of the LDAEP. Moreover, systematic reviews found that ECT has consistently been reported to decrease levels of inflammatory biomarkers, tumor necrosis factor alpha (TNF-*α*) and interleukin-6 (IL-6) (83, 84). Notably, one study demonstrated that the LDAEP was negatively correlated with TNF-*α* (85). This could suggest that a reduction in TNF-*α* might contribute to the LDAEP increase seen after ECT. Given these multifaceted biological interactions, further research is indeed warranted to unravel the complexities of ECT’s impact on the LDAEP and underlying neurobiological mechanisms.

In terms of cognitive performance, our study found that the pre-ECT LDAEP correlated with baseline RBANS total scores. However, change in LDAEP was not associated with changes in cognitive performance post-ECT. While the reasons for these findings warrant further investigation, our preliminary data does suggest a link between LDAEP and cognitive performance metrics.

Finally, it is crucial to acknowledge the limitations inherent in this pilot study. The small sample size of nine participants may not fully represent the broader patient population; these findings must be regarded as exploratory. The impact of the patients’ ongoing psychotropic medication on the LDAEP results also cannot be overlooked. Unlike conventional LDAEP studies that employ EEG, this study utilized MEG, which might affect comparability with existing literature. Despite these limitations, this study provides meaningful insights into the changes in LDAEP following ECT and signals the importance of conducting larger-scale, more controlled research to elucidate these preliminary observations.

## 5 CONCLUSION

Contrary to our initial hypothesis, ECT paradoxically led to an increase in LDAEP, implying a reduction in serotonergic activity. Given the complex roles ECT plays in the brain’s neurochemistry and the multi-faceted nature of LDAEP as a biological marker, our results might not signal a straightforward suppression of serotonin. They could reflect compensatory adjustments in serotonergic receptor activity or broader changes encompassing other neurotransmitter systems, neuroplasticity, and neuro-immune interactions. This unexpected outcome opens avenues for multiple lines of inquiry: the intricate interplay between ECT and LDAEP, and how they might influence 1) the activity of serotonergic, dopaminergic, glutamatergic, and GABAergic receptors and transporters; 2) neuroplasticity and BDNF levels in the temporal cortex; and 3) levels of the pro-inflammatory cytokine TNF-*α*.L ooking forward, further investigation is needed to validate the LDAEP as a biomarker of serotonergic neurotransmission and to elucidate ECT’s effect on serotonergic activity in the human brain.

## Data Availability

All data produced in the present study are available upon reasonable request to the authors.

